# Long-term dynamic changes of choroidal thickness and choroidal vascular index in patients with different types of retinal vein occlusion after intravitreal Ranibizumab

**DOI:** 10.1101/2025.08.01.25332734

**Authors:** Yuanyuan Qi, Jiayang Xu, Daniel Hillarion Scotland, Chao Zhang

**Author notes:** Corresponding Author: Yuanyuan Qi.

## Abstract

**Purpose:** To observe the long-term dynamic changes of choroidal thickness and choroidal vascularity index (CVI) in patients with different types of retinal vein occlusion (RVO) after multiple intravitreal injections of Ranibizumab.

**Methods:** A retrospective cohort study. A total of 70 patients (70 eyes) were treated with intravitreal injection of Ranibizumab from Jan/2022 to Mar/2023 in our hospital. There were 25 cases (25 eyes) of central retinal vein occlusion (CRVO), 45 cases (45 eyes) of BRVO, 10 cases (10 eyes) of DRT, 25 cases (25 eyes) of CME and 35 cases (35 eyes) of MIX. 3+PRN therapy was used. OCT was used to obtain images of the macular region, EDI-OCT was used to obtain clear images of the choroid. Central retina thickness (CRT), subfoveal choroidal thickness (SFCT), the nasal choroidal thickness at 1.5mm of macula (CT N1.5mm), the temporal choroidal thickness at 1.5mm of macula (CT T1.5mm) were measured. Mean macular thickness (CT mean) was calculated. Image J software for binarization processing of choroidal images was used to analyze luminal area (LA), stromal area (SA) and total choroidal area (TCA), and choroidal vascularity index (CVI) was calculated. All above indexes were recorded at baseline, 1 month after each injection, at least 6 months after last injection, and the changes of choroidal indexes in patients with different types of RVO or RVO-ME were compared, and explore the significances of choroidal thickness and CVI as evaluative indicators to predict the therapeutic effects of RVO.

**Results:** 1) BCVA in CRVO group was significantly lower than that in BRVO group, and CRT was significantly higher than that in BRVO group (*P* <0.01). SFCT in CRVO group was higher than that in BRVO group, LA and TCA in CRVO group were lower than those in BRVO group, all the differences were statistically significant (*P* ≤0.05). 2) The CRT, CT of CME group were lower than those of DRT and mixed type, and the differences were statistically significant (*P* ≤0.05). 3) There was no statistically significant difference in CVI between CRVO and BRVO, and no statistically significant difference in CVI between the three different types of macular edema. 4) Regardless of the type of RVO of RVO-ME, BCVA and CRT were significantly improved after multiple injections, and the differences were statistically significant (*P* <0.01). CT, LA, SA, TCA and CVI all decreased gradually after the 1^st^ and 2^nd^ injection compared with baseline, with statistical significance (P <0.01), and remained relatively stable after the 2^nd^ injection. 5) BCVA was further improved compared with the last injection, and CRT, CT, LA, TCA, CVI remained relatively stable with the last injection. There were no statistically significant changes of CVI between the acute stage and the stable stage. 6) CT at baseline was positively correlated with BCVA (LogMAR) and CRT, while CVI at baseline had no correlation with BCVA (LogMAR) and CRT. 7) The CT at baseline was positively correlated with the improvement of BCVA and CRT after treatment, while the CVI at baseline was not correlated with the improvement of BCVA, but was negatively correlated with the improvement of CRT. 8) CVI changes were more pronounced in men compared to women and BRVO was more likely to cause CVI changes than CRVO.

**Conclusions:** RVO not only affects the structure of the retina, but also has certain effects on choroid morphology and blood perfusion. The choroid thickness in the CRVO group was larger than that in the BRVO group. CVI did not significantly differ across RVO or RVO-ME subtypes. After intravitreal Ranibizumab of RVO, choroidal thickness, LA, SA, TCA, and CVI all decreased and remained relatively stable, and choroidal vessels were further remodeled. CVI has no significant correlations with age, visual acuity, type of macular edema and CRT. CVI is a reliable indicator for evaluating choroidal blood perfusion. Choroid thickness and CVI at baseline could be used as predictors of BCVA and CRT improvement after anti-VEGF treatment.

## Introduction

Retinal Vein Occlusion (RVO) is the second most common retinal disease in middle-aged and elderly people that severely affects vision, following diabetic retinopathy. It often leads to vision loss or metamorphopsia due to secondary macular edema. The choroid, composed of stroma and multiple layers of blood vessels, supplies the outer layers of the retina and is an important intraocular structure with a large blood flow. In recent years, an increasing number of studies have found that the choroid is associated with the development of many retinal diseases. Therefore, choroidal imaging for the study of the pathogenesis of various retinal diseases has garnered increasing attention from scholars [1].

Clinically, there are various methods for choroidal imaging. Traditional time-domain OCT is unable to achieve clear choroidal imaging. Currently, spectral-domain OCT with Enhanced Depth Imaging (EDI-OCT) is widely used to present clearer choroidal images for various analyses [2]. Most studies analyze the choroid by assessing the subfoveal choroidal thickness (SFCT). However, since SFCT is influenced by many factors and shows significant variability among different populations [3], many scholars have begun to explore the use of the choroidal vascularity index (CVI) as a research indicator for choroidal observation [4]. The CVI can reflect changes in the vascular components of the choroid in different diseases and the therapeutic effects of these diseases. The CVI is the ratio of the luminal area (LA) of the choroidal vessels to the total choroidal area (TCA). It is obtained using image processing software, with the most commonly used tool being the Niblack automatic local thresholding function in Image J software to binarize the choroidal images, followed by relevant calculations to derive the CVI [5].

A variety of diseases can cause pathological changes in the structure of the choroid. For example, central serous chorioretinopathy (CSC) and polypoidal choroidal vasculopathy (PCV) are both included in the spectrum of thick choroid diseases and have become a hot topic in choroidal research [6]. Currently, the CVI has been relatively widely applied in the study of PCV, AMD, CSC, and DR. Studies have found that during the acute phase of CSC, the CVI of the affected eye significantly increases, indicating a state of high choroidal vascular perfusion. However, after the acute phase subsides, the CVI of the affected eye returns to normal [7]. Other scholars have found that in CSC patients with choroidal neovascularization (CNV), the CVI and SFCT are significantly lower than those in CSC patients without CNV, suggesting that choroidal ischemia may be a potential factor promoting the development of CNV [8]. Multiple studies have shown that the CVI in PCV patients is significantly lower than that in healthy individuals [9–10]. Some scholars have used the CVI to classify PCV into two subtypes: high-permeability type (high CVI and high SFCT) and low-permeability type (low CVI and low SFCT). Research on the CVI has expanded the understanding of the pathogenesis of PCV [11]. There are also relevant studies on neovascular AMD (nAMD). Some scholars have found that the CVI in AMD-affected eyes is significantly lower than that in normal individuals, suggesting that choroidal ischemia may be the cause of CNV formation secondary to nAMD [12]. Other scholars have used the CVI to observe the therapeutic effects of photodynamic therapy (PDT) on nAMD and found that after PDT treatment, both the CVI and SFCT significantly decreased [13]. In addition to extensive research on thick choroid diseases, studies on the choroid in diabetic retinopathy patients have also become popular in recent years. Some scholars have found that the CVI in DR patients is significantly lower than that in healthy populations, and it further decreases with the progression of DR. It is believed that the progression of diabetes affects the choroidal blood supply, which in turn affects the blood supply to the outer retina.

Persistent choroidal ischemia may be one of the important reasons for the progression of diabetic retinopathy [14]. In addition to the diseases mentioned above that have been extensively studied for their effects on the choroid, research on other retinal diseases, such as retinal vein occlusion, uveitis, and retinal degenerative diseases, has also gradually increased, although the number of related reports is still limited [15–17]. From the above reports, it is evident that many retinal diseases not only affect the retina but also involve the choroid. Changes in choroidal CVI and SFCT can serve as non-invasive tools to monitor changes in choroidal structure in retinal diseases, assess the therapeutic effects of diseases, and help understand the pathogenesis of diseases. Intravitreal injection of anti-vascular endothelial growth factor (anti-VEGF) agents has become the first-line treatment for macular edema caused by RVO. However, repeated intravitreal injections are often required in clinical practice to achieve therapeutic effects. Previous studies have mainly focused on the impact of common thick choroid diseases or diabetic retinopathy on choroidal indicators and the changes in choroidal structure after retinal laser treatment for these retinal diseases. In contrast, reports on choroidal observation in RVO patients are relatively scarce, especially regarding the changes in CVI and the therapeutic response after multiple anti-VEGF treatments in patients with different types of RVO and RVO-ME. In view of the relative limitations of the above studies, this study adopts a retrospective study method. We observed the changes in choroidal thickness and CVI after multiple anti-VEGF treatments in acute RVO patients, the changes in choroidal thickness and CVI in RVO patients after at least six months of treatment cessation following edema resolution, and the comparison of choroidal thickness and CVI at baseline and their response to multiple anti-VEGF treatments in patients with different types of RVO and RVO-ME. The aim is to explore the dynamic changes in choroidal indicators during the acute and stable phases of RVO and to investigate the potential of choroidal thickness and CVI as predictive indicators for assessing the therapeutic effects in patients with different types of RVO and RVO-ME. This study also aims to provide more comprehensive reference data for the clinical research of CVI.

## Materials and Methods

### Research Subjects

This was a retrospective cohort study. A total of 70 eyes from 70 patients with Retinal Vein Occlusion (RVO) who underwent intravitreal injection of ranibizumab (0.5mg/0.05ml) at our hospital’s intravitreal injection center from January 2022 to June 2023 were selected. Data were accessed for research purposes starting on November 4, 2023.The cohort included 34 males (34 eyes) and 36 females (36 eyes), with a mean age of 58.25 ± 10.63 years. There were 25 eyes with Central Retinal Vein Occlusion (CRVO) and 45 eyes with Branch Retinal Vein Occlusion (BRVO). All enrolled cases were treated with intravitreal injection in one eye and followed up with a 3+PRN (pro re nata) treatment regimen, which involved monthly injections for three consecutive months, followed by monthly reexamination to determine the timing and regimen for subsequent injections. The research protocol received approval from the Ethics Committee of the Dalian Third People’s Hospital (Approval Number: 2023-145-001). This study adhered to the principles outlined in the Declaration of Helsinki. In accordance with the requirements set forth by the ethics committee, the waiver of patient informed consent was granted. The authors did not have access to any information that could identify individual participants during or after the data collection process. Trial registration: ChiCTR, ChiCTR2400090054.

### Inclusion Criteria

(1) Clinically diagnosed with unilateral RVO, presenting with decreased vision due to macular edema, requiring intravitreal injection of anti-VEGF agents. (2) Central foveal thickness measured by OCT greater than 300μm at the initial visit. (3) No prior history of intravitreal injections or retinal laser photocoagulation. (4) No prior history of any intraocular surgery.

### Exclusion Criteria

(1) Presence of macular edema caused by other etiologies besides RVO (e.g., diabetic macular edema (DME), neovascular AMD (nAMD), vitreomacular traction, uveitis, etc.). (2) Presence of other ocular diseases that may affect vision. (3) Refractive error with spherical equivalent ≥±6.0D. (4) Axial length of the eye >26mm or <22mm. (5) Disease duration exceeding three months. (5) Presence of ocular or severe systemic diseases that may affect choroidal blood flow (e.g., diabetes). (6) Unclear choroidal imaging on EDI-OCT (e.g., due to vitreous hemorrhage, severe cataract, or in some acute RVO cases where severe retinal hemorrhage or high macular edema obscures the underlying choroidal signal).

### Method of Examination

General information including age, gender, and systemic diseases was recorded. Ocular examinations included best-corrected visual acuity (BCVA), intraocular pressure (IOP), slit-lamp examination, funduscopy, and fundus photography. Optical coherence tomography (OCT) was performed using SD-OCT (Heidelberg) to confirm the diagnosis and classification of macular edema, and EDI-OCT mode was used for choroidal imaging. Postoperative follow-up was conducted to monitor and assess treatment efficacy to formulate subsequent treatment plans. BCVA was measured using a standard logarithmic visual acuity chart and converted to LogMAR (Logarithm of the minimum angle of resolution) for statistical analysis. Data for all the above indicators were recorded at baseline before surgery and one month after each injection (as short-term observation indicators), as well as at least six months after edema resolution and stabilization when injections were discontinued (as long-term observation indicators), to observe the dynamic changes in choroidal indicators after multiple intravitreal injections of ranibizumab during the acute phase of RVO and at least six months after stabilization following edema resolution.

### Macular Edema Classification

Based on OCT findings, macular edema was classified into three types according to morphological characteristics. According to the classification of macular edema types, there were 10 eyes with DRT, 25 eyes with CME, and 35 eyes with MIX. Type I: Diffuse retinal thickening (DRT), characterized by diffuse sponge-like retinal edema in the macular area with uniform reduction of retinal reflectivity (Figure 1A).Type II: Cystoid macular edema (CME), characterized by one or more low-reflective cystic cavities in the macular retina (Figure 1B).Type III: Mixed type (MIX): Serous retinal detachment (SRD) with CME, defined by the presence of a low-reflective space (SRD) between the retinal pigment epithelium and the neurosensory layer, along with low-reflective cystic cavities within the macular retina (Figure 1C).

**Figure 1.**
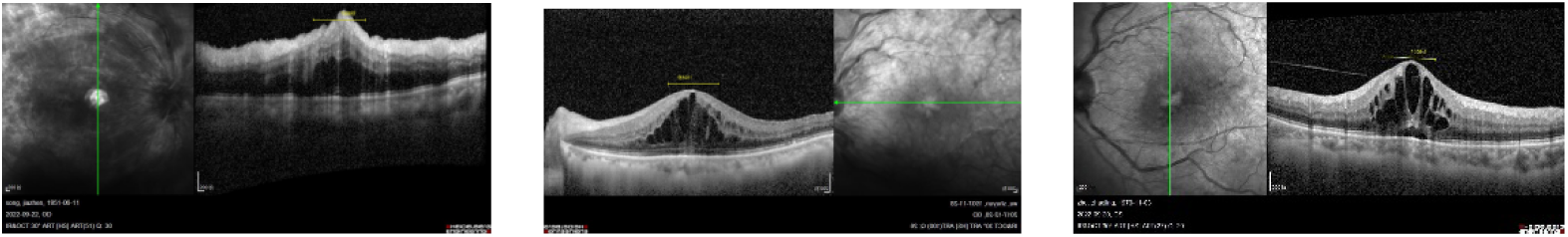
A: Diffuse retinal thickening (DRT). Figure 1B: Cystoid macular edema (CME). Figure 1C: Serous retinal detachment (SRD) with CME.

### Measurement and Recording

All OCT examinations and measurements of choroidal thickness were performed by the same experienced OCT operator. The central retinal thickness (CRT) of all patients was recorded using the embedded tool. The choroidal thickness was measured using the built-in caliper of the instrument software, from the outer edge of the retinal pigment epithelium’s high-reflective layer to the chorioscleral border.

Three measurements were taken in each eye: at the fovea (SFCT), 1500 μm nasal to the fovea (CT N1.5mm), and 1500 μm temporal to the fovea (CT T1.5mm). The average of these three measurements (CT Mean) was calculated as the mean choroidal thickness in the macular center area. The same experienced individual then processed the images and calculated the LA, SA, TCA, and CVI values within a 1500μm radius centered on the fovea using Image J software.

### Image J Software Image Processing Method **[13]**

The same image processor handled all EDI-OCT images. The choroidal vascular areas were outlined using the polygon tool and recorded via the selection manager. The pixel-to-size correspondence was removed, and the image was sampled multiple times (with a sampling range of 13px × 13px) in the center of the vascular lumens to measure the grayscale values at four sampling points. The average grayscale value was adjusted to make the darker areas of the image appear blacker. The image type was then changed to 8-bit and adjusted using the Niblack automatic local threshold. The choroidal region in the OCT scan was binarized, and each binarized image was converted to RGB format. The lumen areas were highlighted using the color threshold tool, and the total choroidal area, choroidal lumen area, and choroidal stroma area within the 1500μm macular region were calculated. Bright pixels were defined as the choroidal stroma, while dark pixels were defined as the choroidal vascular lumen areas. (Figure 2).

**Figure 2.**
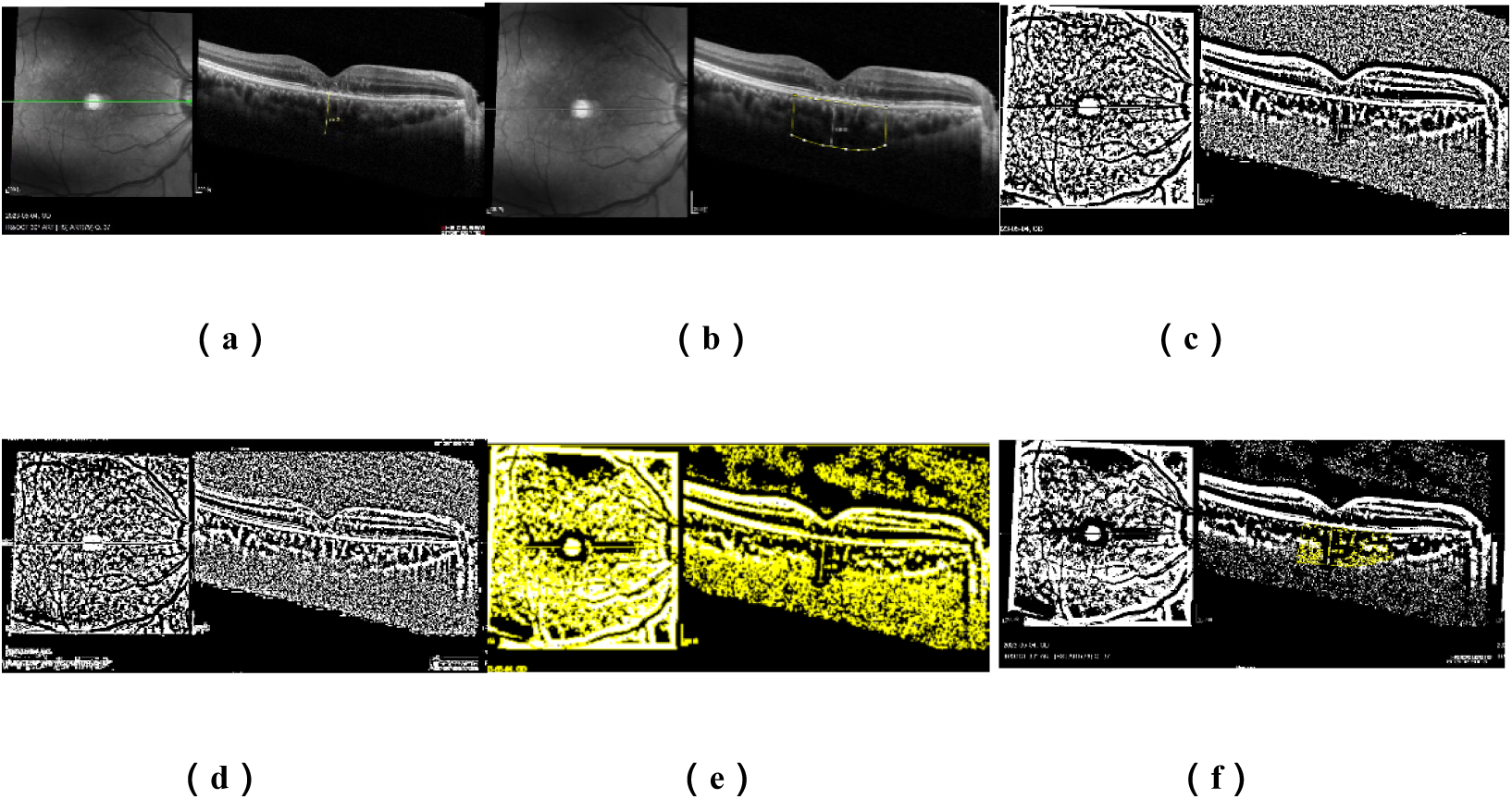
Image Binarization: (a) SD-OCT image (EDI mode); (b) Polygonal selection of the choroid under the 1500μm horizontal line centered on the macular fovea; (c) Image binarization using the Niblack method; (d) Minor adjustment of the polygonal points for better visualization at the chorioscleral junction; (e) Color thresholding to highlight the vascular regions; (f) The final result with the lumen area measured under the 1500μm horizontal line centered on the fovea superimposed on the SD-OCT image.

### Treatment

After thoroughly cleaning the conjunctival sac, topical application of proparacaine hydrochloride (Alcon) was administered to the eye, one drop every 5 minutes for a total of three times. The area was then disinfected and draped. The conjunctival sac was soaked with 0.05% povidone-iodine (Li Erkang, Shandong) for 1 minute and 30 seconds. Using a 30G needle, the drug (ranibizumab 0.5mg/0.05ml) was injected into the vitreous cavity 3.5-4mm posterior to the limbus. Post-injection light perception was assessed, and the procedure was concluded. All patients were prescribed levofloxacin hydrochloride eye drops (Cravit, Santen), to be used four times daily for seven consecutive days following the procedure.

### Statistical Analysis

Statistical analysis was performed using SPSS 22.0 software. Data with a normal distribution were expressed as mean ± standard deviation. For comparisons between different types of vein occlusion, independent t-tests were used for continuous variables, and chi-square (χ²) tests were used for categorical variables. Paired t-tests were used to compare indicators between the affected eyes and the fellow eyes. For comparisons among multiple groups of patients with different types of macular edema, one-way analysis of variance (ANOVA) was used for continuous variables, and χ² tests were used for categorical variables. For comparisons of continuous variables among multiple groups after injection, covariance analysis was used with baseline correction. Repeated-measures ANOVA was used for within-group comparisons of indicators at different time points before and after surgery, with multiple comparisons conducted using the LSD-t test. Pearson correlation analysis was used to assess the correlation between continuous variables. Multiple linear regression analysis was used for regression analysis. A P-value of ≤0.05 was considered statistically significant.

## Results

### 1 Comparison of indicators at baseline

#### 1.1 Comparison of indicators among different types of RVO at baseline

Table 1 presents the basic information of patients with different types of RVO. There were no significant differences in mean age, gender, or affected eye between the CRVO group and the BRVO group (Table 1).

**Table 1.** Basic information of patients with different types of RVO.

| Index | CRVO (n=25) | BRVO (n=45) | t/x<sup>2</sup> | P |
| --- | --- | --- | --- | --- |
| Age | 57.72±11.66 | 58.78± 9.59 | -0.41 | 0.68 |
| Sex male (%) | 7(28%) | 27(60%) | 3.71 | 0.054 |
| Eye right (%) | 11 (44%) | 30 (66.67%) | 3.40 | 0.065 |

Note: CRVO: central retinal vein occlusion; BRVO: branch retinal vein occlusion.

Table 2 presents the comparison of baseline indicators in patients with different types of RVO. The BCVA in the CRVO group was significantly lower than that in BRVO group, and the CRT was significantly higher than that in BRVO group, with statistically significant differences (P < 0.05). The SFCT in the CRVO group was larger than that in BRVO group, while the LA and TCA were smaller than those in BRVO group, with statistically significant differences (P ≤ 0.05). There was no statistically significant difference in CVI values between the two groups (Table 2).

**Table 2.**
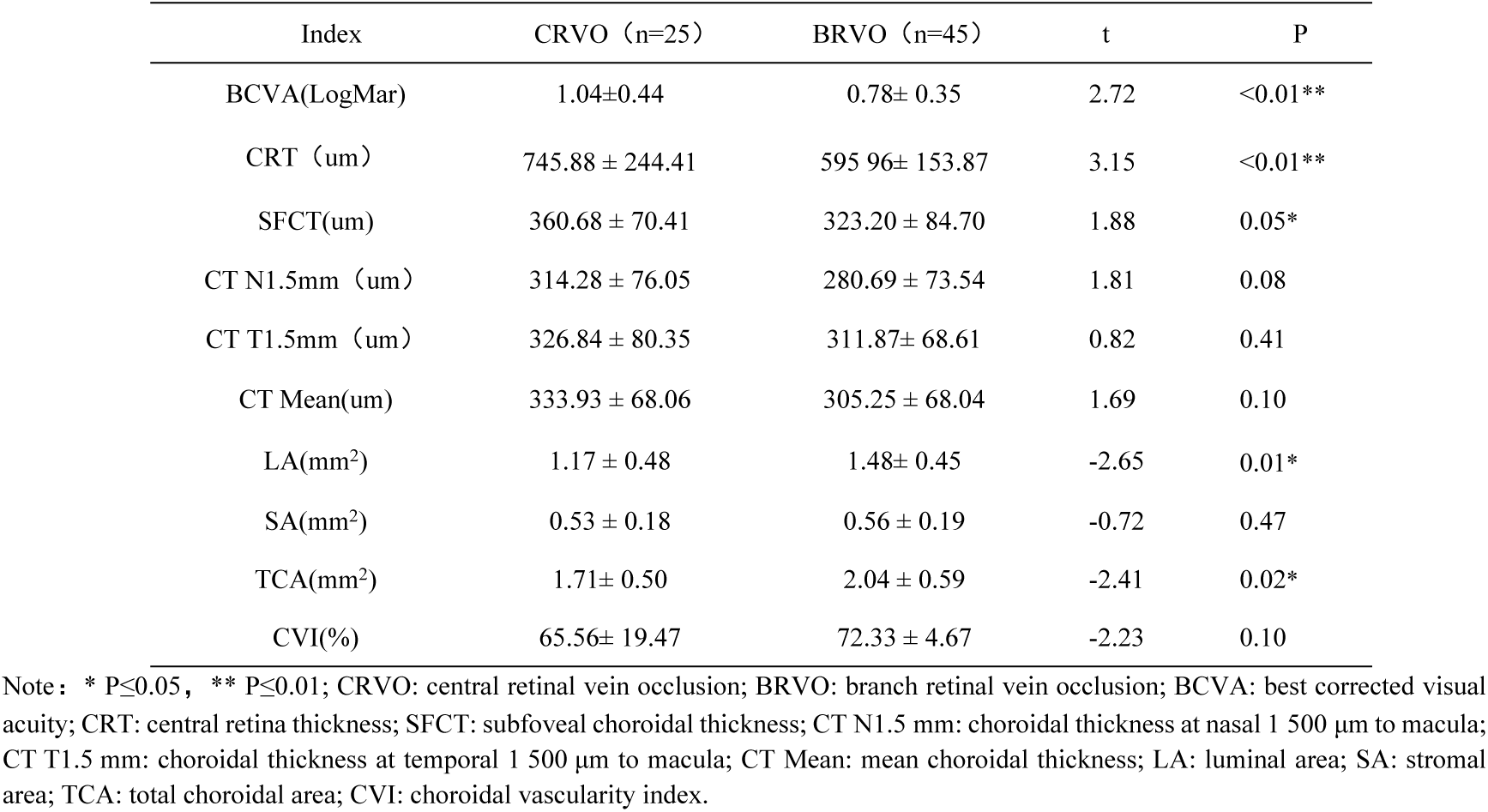
Comparison of baseline indicators in patients with different types of RVO.

| Index | CRVO (n=25) | BRVO (n=45) | t | P |
| --- | --- | --- | --- | --- |
| BCVA(LogMar) | 1.04±0.44 | 0.78± 0.35 | 2.72 | <0.01** |
| CRT (um) | 745.88 ± 244.41 | 595.96± 153.87 | 3.15 | <0.01** |
| SFCT(um) | 360.68 ± 70.41 | 323.20 ± 84.70 | 1.88 | 0.05* |
| CT N1.5mm (um) | 314.28 ± 76.05 | 280.69 ± 73.54 | 1.81 | 0.08 |
| CT T1.5mm (um) | 326.84 ± 80.35 | 311.87± 68.61 | 0.82 | 0.41 |
| CT Mean(um) | 333.93 ± 68.06 | 305.25 ± 68.04 | 1.69 | 0.10 |
| LA(mm<sup>2</sup>) | 1.17 ± 0.48 | 1.48± 0.45 | -2.65 | 0.01* |
| SA(mm<sup>2</sup>) | 0.53 ± 0.18 | 0.56 ± 0.19 | -0.72 | 0.47 |
| TCA(mm<sup>2</sup>) | 1.71± 0.50 | 2.04 ± 0.59 | -2.41 | 0.02* |
| CVI(%) | 65.56± 19.47 | 72.33 ± 4.67 | -2.23 | 0.10 |

Note: \* $P \leq 0.05$ , \*\* $P \leq 0.01$ ; CRVO: central retinal vein occlusion; BRVO: branch retinal vein occlusion; BCVA: best corrected visual acuity; CRT: central retina thickness; SFCT: subfoveal choroidal thickness; CT N1.5 mm: choroidal thickness at nasal 1 500 μm to macula; CT T1.5 mm: choroidal thickness at temporal 1 500 μm to macula; CT Mean: mean choroidal thickness; LA: luminal area; SA: stromal area; TCA: total choroidal area; CVI: choroidal vascularity index.

#### 1.2 Comparison of indicators among different types of macular edema at baseline

Table 3 presents the basic information of patients with different types of macular edema. At baseline, there were no significant differences in mean age, gender, or affected eye among patients with different types of macular edema (Table 3).

**Table 3.** Basic information of patients with different types of macular edema.

| Index | DRT (n=10) | CME (n=25) | MIX (n=35) | F/$\chi^2$ | P |
| --- | --- | --- | --- | --- | --- |
| Age | 57±6.68 | 61.04±10.34 | 56.91±10.96 | 1.288 | 0.28 |
| Sex male (%) | 6(60%) | 10(40%) | 18(51.4%) | 1.373 | 0.50 |
| Eye right (%) | 8(80%) | 12(48%) | 21(60%) | 3.073 | 0.22 |

Note: DRT: Diffuse retinal thickening; CME: Cystoid macular edema; MIX: Serous retinal detachment (SRD) with CME.

Table 4 presents the comparison of baseline indicators in patients with different types of macular edema. The CRT in CME group was lower than that in DRT and mixed types, with statistically significant differences (P < 0.01). The SFCT, CT N1.5mm, and CT mean in CME group were smaller than those in other two groups, with statistically significant differences (P ≤ 0.05). There were no statistically significant differences in CVI among the three groups (Table 4).

**Table 4.** Comparison of baseline indicators in patients with different types of macular edema.

| Index | DRT (n=10) | CME (n=25) | MIX (n=35) | F | p |
| --- | --- | --- | --- | --- | --- |
| BCVA(LogMar) | 0.73 ± 0.40 | 0.88 ± 0.31 | 0.92 ± 0.42 | 0.872 | 0.42 |
| CRT (um) | 658.20 ± 168.17<sup>ab</sup> | 557.32 ± 161.26<sup>ac</sup> | 715.71 ± 216.39<sup>b</sup> | 4.961 | 0.01* |
| SFCT(um) | 370 ± 86.08<sup>a</sup> | 299.72 ± 70.4<sup>b</sup> | 353.37 ± 79.81<sup>a</sup> | 4.582 | 0.01* |
| CT N1.5mm (um) | 335.10 ± 67.70<sup>a</sup> | 268.32 ± 87.25<sup>b</sup> | 297.97 ± 63.51<sup>a</sup> | 3.137 | 0.05* |
| CT T1.5mm (um) | 360.50 ± 115.50 | 300.40 ± 67.05 | 316.86 ± 57.37 | 2.544 | 0.09 |
| CT Mean(um) | 355.20 ± 85.59<sup>a</sup> | 289.48 ± 68.21<sup>b</sup> | 322.73 ± 58.40<sup>a</sup> | 3.939 | 0.02* |
| LA(mm<sup>2</sup>) | 1.49 ± 0.30 | 1.31 ± 0.45 | 1.38 ± 0.55 | 0.509 | 0.60 |
| SA(mm<sup>2</sup>) | 0.55 ± 0.14 | 0.51 ± 0.18 | 0.58 ± 0.20 | 1.022 | 0.37 |
| TCA(mm<sup>2</sup>) | 2.03 ± 0.33 | 1.82 ± 0.60 | 1.96 ± 0.63 | 0.642 | 0.53 |
| CVI(%) | 72.97 ± 6.36 | 71.71 ± 3.52 | 67.77 ± 16.98 | 1.073 | 0.35 |

Note: \* $P \leq 0.05$ , \*\* $P \leq 0.01$ ; different letter superscripts without duplication indicate that multiple comparison differences are statistically significant. DRT: Diffuse retinal thickening; CME: Cystoid macular edema; MIX: Serous retinal detachment (SRD) with CME; BCVA: best corrected visual acuity; CRT: central retina thickness; SFCT: subfoveal choroidal thickness; CT N1.5 mm: choroidal thickness at nasal 1 500 μm to macula; CT T1.5 mm: choroidal thickness at temporal 1 500 μm to macula; CT Mean: mean choroidal thickness; LA: luminal area; SA: stromal area; TCA: total choroidal area; CVI: choroidal vascularity index.

### 2 Dynamic changes in indicators of different types of RVO eyes after multiple injections

Table 5 presents indexes in different types of RVO in stable stage and acute stage. Comparison of the levels of indicators at different time points during the acute phase and after at least six months of treatment cessation in eyes that underwent multiple intravitreal injections (stable phase). In RVO patients, whether CRVO or BRVO, the BCVA and CRT of the affected eyes were significantly improved after multiple injections, with statistically significant differences (P < 0.01). After stopping the injections for six months, the BCVA further improved compared to the last injection, while the CRT remained relatively stable compared to the last injection. Choroidal thickness (SFCT, CT N1.5mm, CT T1.5mm, CT Mean) gradually decreased after the first injection compared to baseline, with statistically significant differences (P < 0.01), and remained relatively stable after the second injection until at least six months after the last injection. The LA, SA, and TCA of CRVO and BRVO gradually decreased after anti-VEGF injection compared to baseline, with statistically significant differences (P < 0.01), and the downward trend of LA and TCA continued until at least six months after the last injection. The CVI in CRVO continuously decreased after injection, but the change was not statistically significant. In the BRVO group, the CVI significantly decreased after the second injection and remained stable thereafter, still maintaining stability at least six months after the last injection, with statistically significant differences (P < 0.01) (Table 5).

**Table 5.**
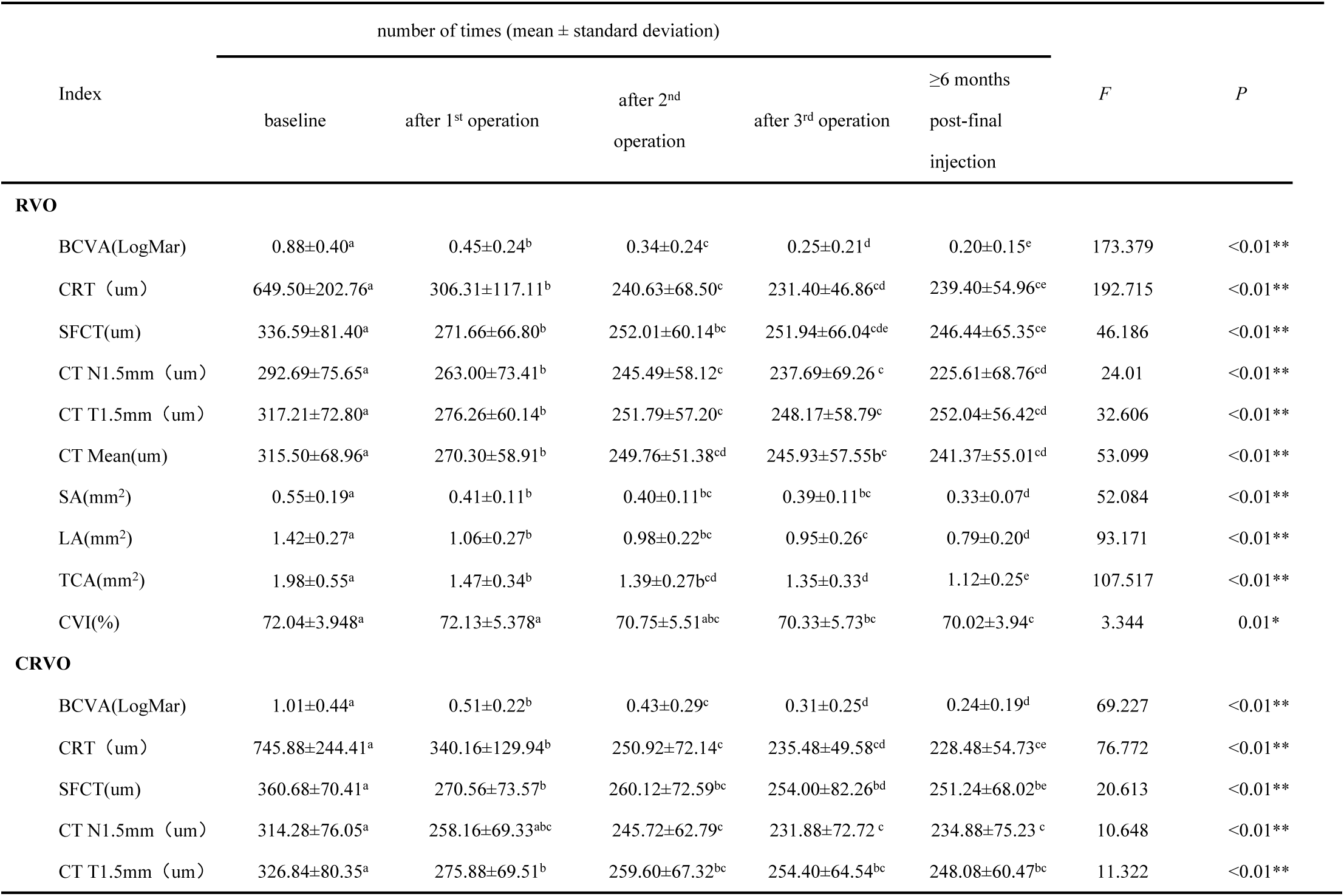
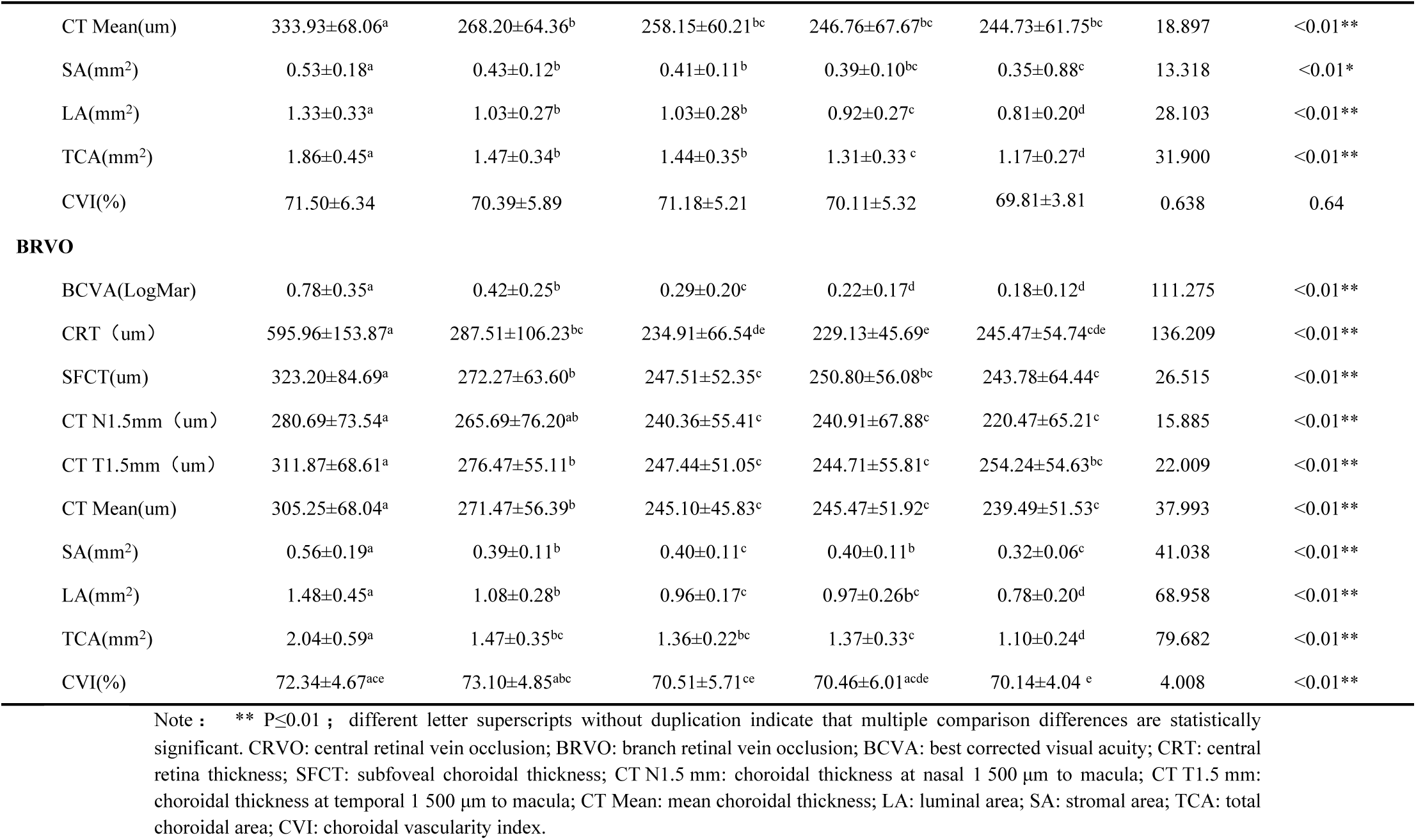
Indexes in different types of RVO in stable stage and acute stage.

### 3 Dynamic changes in indicators of different types of macular edema eyes after multiple injections

Table 6 presents indexes in different types of macular edema compared with stable stage and acute stage. Comparison of the levels of indicators at different time points during the acute phase and after at least six months of treatment cessation in eyes that underwent multiple intravitreal injections (stable phase). In RVO-ME patients, regardless of the type of edema, the BCVA and CRT of the affected eyes were significantly improved after multiple injections, with statistically significant differences (P < 0.01), and remained stable for at least six months after the last injection.The choroidal thickness (SFCT, CT N1.5mm, CT T1.5mm, CT Mean) of all three types of edema decreased after the first injection compared to baseline, with statistically significant differences (P < 0.01), and remained relatively stable after the second injection until at least six months after the last injection. The LA, SA, and TCA of all three types of edema decreased after the first injection compared to baseline, with the downward trend continuing until at least six months after the last injection, showing statistically significant differences (P < 0.01).There were no statistically significant differences in the changes of CVI between the acute phase and the stable phase for all three types of edema (Table 6).

**Table 6.**
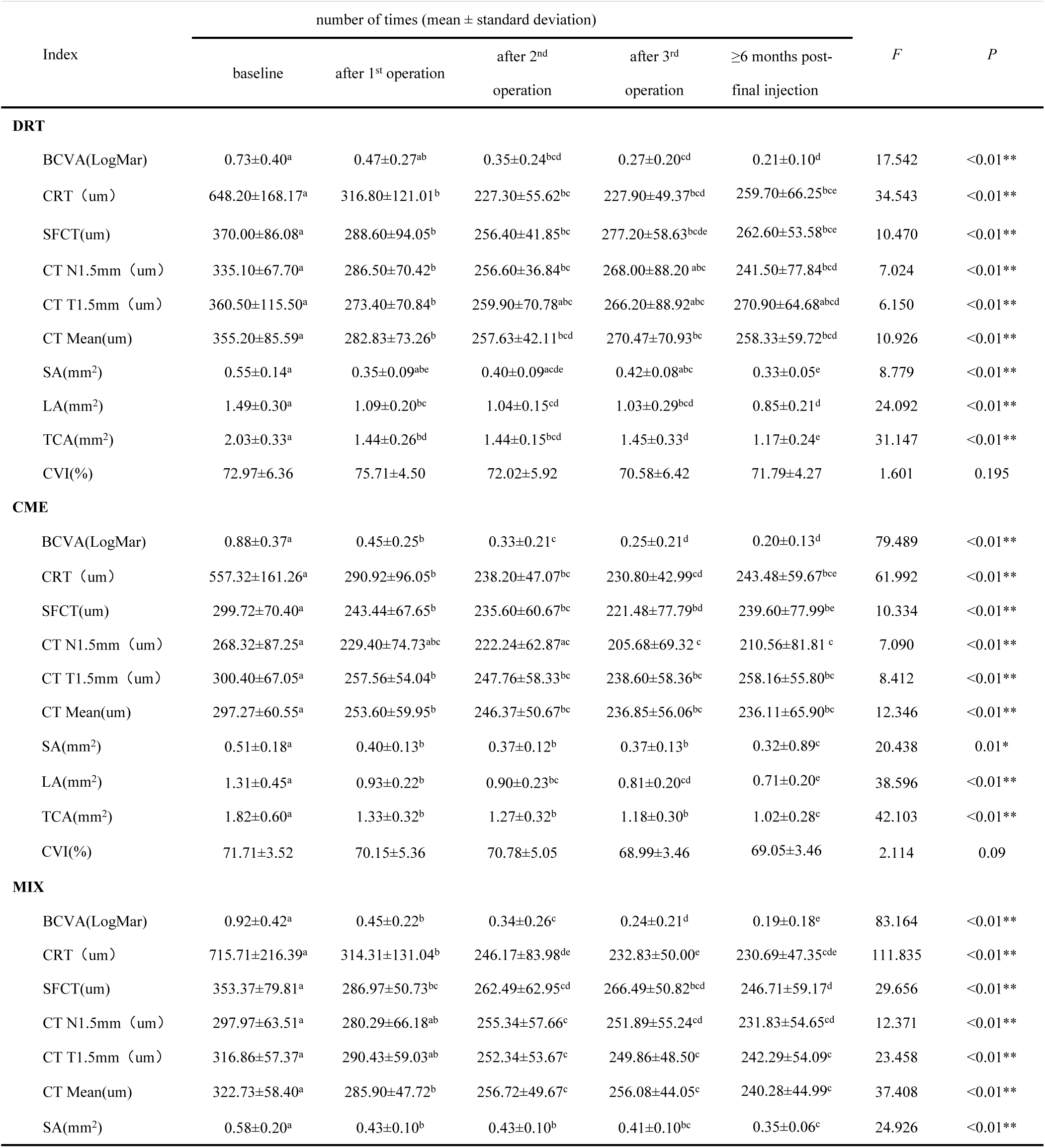
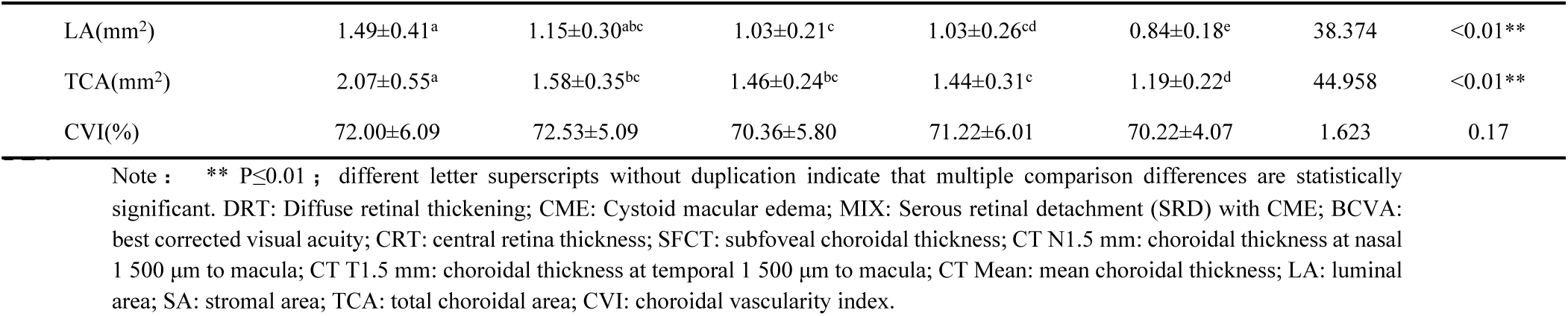
Indexes in different types of macular edema compared with stable stage and acute stage.

### 4 Correlation Analysis

#### 4.1 Correlation between choroidal indicators at baseline and baseline visual acuity and central retinal thickness

Table 7 presents correlation of baseline visual acuity, CRT, and choroid index. The choroidal thickness at baseline was positively correlated with baseline BCVA (LogMAR) and baseline CRT. CVI at baseline showed no significant correlation with baseline BCVA (LogMAR) or baseline CRT (Table 7).

**Table 7.** Correlation of baseline visual acuity, CRT, and choroid index.

| Index | baseline BCVA(LogMar) | baseline CRT |
| --- | --- | --- |
| baseline SFCT | 0.299** ( <0.01) | 0.386** ( <0.01 ) |
| baseline CT Mean | 0.304**(<0.01) | 0.358** ( <0.01) |
| baseline CVI | -0.088(0.29) | -0.154*(0.054) |

Note: the data in the table is pearson correlation coefficient (P-value). BCVA: best corrected visual acuity; CRT: central retina thickness; SFCT: subfoveal choroidal thickness; CT Mean: mean choroidal thickness; CVI: choroidal vascularity index.

#### 4.2 Correlation between baseline choroidal indicators and improvement in BCVA and CRT after treatment

Table 8 presents the correlation between choroidal indices at baseline and improvement of BCVA and CRT after treatment. Baseline choroidal thickness was not correlated with post-treatment BCVA or post-treatment CRT but was positively correlated with the improvement in BCVA and the improvement in CRT after treatment. Baseline CVI was not correlated with the improvement in BCVA after treatment but was negatively correlated with the improvement in CRT after treatment. (Table 8)

**Table 8.** Correlation between choroidal indices at baseline and improvement of BCVA and CRT after treatment.

| Index | BCVA<br>(after treatment) | CRT<br>(after treatment) | BCVA<br>(improvement) | CRT<br>(improvement) |
| --- | --- | --- | --- | --- |
| baseline SFCT | 0.082 ( 0.31 ) | 0.025 ( 0.76 ) | 0.274** ( <0.01 ) | 0.363** (<0.01) |
| baseline CT Mean | 0.046 (0.57) | -0.006 ( 0.94) | 0.305** ( <0.01 ) | 0.344** (<0.01) |
| baseline CVI | -0.129 (0.11) | 0.097 (0.23) | - 0.004 (0.96) | - 0.171* (0.03) |

Note: the data in the table is pearson correlation coefficient (P-value). BCVA: best corrected visual acuity; CRT: central retina thickness; SFCT: subfoveal choroidal thickness; CT Mean: mean choroidal thickness; CVI: choroidal vascularity index.

### Multivariate Regression Analysis of Changes in Choroidal Indicators

Table 9 presents the Multivariate regression analysis of mean choroidal thickness. Gender, age, type of RVO, type of RVO macular edema, baseline visual acuity were not significantly correlated with changes in mean choroidal thickness or choroidal vascularity index (CVI). Baseline CRT was negatively correlated with changes in mean choroidal thickness (Table 9). Table 10 presents the multivariate regression analysis of choroidal vascular index. Compared to females, males were more likely to experience changes in CVI. Compared to CRVO, BRVO was more likely to cause changes in CVI. However, age, type of macular edema, baseline visual acuity, and baseline CRT showed no significant correlation with changes in CVI (Table 10).

**Table 9.** Multivariate regression analysis of mean choroidal thickness.

| Index | Estimate | P | 95.00% |
| --- | --- | --- | --- |
| Sex | -18.020 | 0.19 | 4.942 |
| BRVO | 12.055 | 0.44 | 38.003 |
| Age | -1.345 | 0.06 | -0.198 |
| DRT | 0 |  |  |
| CME | 38.320 | 0.10 | 76.251 |
| MIX | 27.597 | 0.18 | 61.874 |
| Baseline BCVA | -12.201 | 0.82 | 78.038 |
| Baseline CRT | -0.115 | 0.01* | -0.041 |

Note: \*P≤0.05. BCVA: best corrected visual acuity; CRT: central retina thickness; DRT: Diffuse retinal thickening; CME: Cystoid macular edema; MIX: Serous retinal detachment (SRD) with CME.

**Table 10.** Multivariate regression analysis of choroidal vascular index.

| Index | Estimate | P | 95.00% |
| --- | --- | --- | --- |
| Sex male | -8.218 | 0.01* | -3.188 |
| BRVO | -7.718 | 0.03* | -2.034 |
| Age | -0.272 | 0.078 | -0.021 |
| Hypertension | -5.589 | 0.088 | -0.335 |
| DRT | 0 |  |  |
| CME | -6.318 | 0.21 | 1.991 |
| MIX | 1.099 | 0.81 | 8.608 |
| Baseline BCVA | -17.524 | 0.14 | 2.244 |
| Baseline CRT | -0.009 | 0.35 | 0.007 |

Note : \*P≤0.05. BCVA: best corrected visual acuity; CRT: central retina thickness; DRT: Diffuse retinal thickening; CME: Cystoid macular edema; MIX: Serous retinal detachment (SRD) with CME.

## Discussion

Retinal vein occlusion (RVO) is a common retinal vascular disease in middle-aged and elderly people, with a prevalence of 0.52% to 0.77%. It can cause macular edema, intraretinal hemorrhage, retinal ischemia, and other conditions that lead to significant vision loss [18]. Currently, the pathogenesis of RVO is not fully understood. The main risk factors include hypertension, diabetes, atherosclerosis, hyperlipidemia, hyper-viscosity of blood, abnormal coagulation function, and glaucoma, among which hypertension is considered the most important risk factor [18]. Recent studies suggest that the occurrence of RVO is due to various causes leading to endothelial damage of the vascular wall, resulting in retinal circulation disorders. The areas along the retinal veins become ischemic and hypoxic, leading to elevated levels of various cytokines and inflammatory factors, which disrupt the retinal barrier and subsequently cause macular edema. The most important factor involved in the disease process is the VEGF factor. Therefore, intravitreal injection of anti-VEGF drugs is currently considered the first-line treatment for RVO-related macular edema [19].

The choroid is the largest vascular-rich tissue structure in the human eye, with choroidal blood flow accounting for 85% of all ocular blood flow [20]. Therefore, due to changes in hydrostatic pressure and VEGF levels in RVO patients, the choroid may also be affected in addition to retinal lesions. In recent years, researchers have begun to explore the changes in the choroid caused by various retinal diseases.

Optical coherence tomography (OCT) is a non-invasive imaging tool that can be used to detect the structure of the retina and choroid in vivo. The enhanced depth imaging (EDI) mode can display the deeper choroidal layers more clearly. Many scholars have used EDI-OCT to observe changes in choroidal thickness in RVO patients. Hae Min Kang et al. [21] reported that in eyes affected by RVO, the choroidal thickness around the optic disc and under the fovea significantly decreased within 12 months. Muge Coban-Karatas et al. [22] reported that the SFCT in RVO patients was significantly thicker than that in the fellow eyes, and the SFCT significantly decreased after anti-VEGF treatment. Other studies have suggested that there is a significant difference in SFCT between RVO eyes and fellow eyes [23], but the SFCT did not decrease after anti-VEGF treatment [24]. These inconsistent reports regarding choroidal thickness and its changes after treatment in RVO eyes suggest that factors influencing choroidal thickness may not be singular. Studies have reported that the choroid, a highly vascularized tissue, is anatomically and physiologically very similar to the glomeruli. Elevated arterial pressure is a major risk factor for both RVO and renal damage, so elevated blood pressure or renal injury may have some impact on the choroid [25, 26]. We believe that changes in choroidal thickness in retinal diseases are not solely related to the VEGF factor and that other confounding factors must be further explored.

The choroidal vascularity index (CVI) originated from the research of Sonoda et al., who binarized the choroidal images under EDI-OCT, with black representing the luminal area (LA) and white representing the stromal area (SA). The sum of the two is the total choroidal area (TCA), and LA/TCA is used to represent choroidal blood flow perfusion [5]. This method was later improved by Agrawal et al. and named choroidal vascularity index (CVI) [3]. Since CVI is not affected by age, refractive error, axial length, or intraocular pressure, it is considered a relatively effective tool for assessing changes in choroidal vasculature and has been used in the study of various ocular diseases, including CSC, AMD, and DME [27–30].

Although RVO primarily affects the retina, many studies have shown that the hypoxia in RVO can further induce changes in the choroid. Our study aims to explore whether CRVO and BRVO induce changes in choroidal vasculature assessed by CVI, whether there are differences in choroidal thickness and CVI between the two, whether there is a relationship between the choroidal response to retinal vein occlusion in different types of macular edema patients, and the trend of changes in choroidal indicators during the acute and stable phases of RVO.

In our cases, 62.12% of CRVO patients had hypertension, while 43.9% of BRVO patients had hypertension, with a statistically significant difference between the two. Clinically, we have observed that CRVO patients often have systemic diseases such as hypertension, hyperlipidemia, and diabetes, with a high proportion of hypertension. In contrast, BRVO patients are more likely to have atherosclerosis as the main cause, and the proportion of those with systemic diseases is relatively lower than that of CRVO patients [31]. Our cases also show that in both CRVO and BRVO, CME type macular edema is the most common, followed by the mixed type, with DRT type being relatively less common and pure SRD type being even rarer. Since diabetic patients themselves can cause diabetic choroidopathy, we excluded diabetic patients in our study to avoid interference.

We know that the acute onset of RVO leads to elevated intraocular VEGF levels, which in turn cause secondary choroidal edema. Elevated VEGF stimulates the production of nitric oxide, vasodilation, and increased ocular blood flow. Elevated VEGF levels are the main cause of increased vascular permeability and choroidal thickness [32, 33]. The ischemic index and VEGF levels in CRVO eyes are higher than those in BRVO eyes [34]. Therefore, it is speculated that the degree of macular edema and choroidal thickness in CRVO eyes should be greater than those in BRVO eyes. Our study confirmed this speculation. We found that the central retinal thickness, subfoveal choroidal thickness, and mean choroidal thickness in the CRVO group were all thicker than those in the BRVO group, which is consistent with the results reported in previous literature [35, 36]. The elevated VEGF levels can also explain why CRVO eyes have increased central retinal thickness and worse visual acuity. At the same time, we found that there was no significant difference in CVI between CRVO and BRVO, which is consistent with the reports of Loiudice P et al. [37]. This suggests that changes in choroidal vasculature may be unrelated to the type of occlusion.

Previous studies have shown that the SFCT in CRVO eyes is significantly thicker than that in other eyes, and the SFCT significantly decreased two weeks after anti-VEGF treatment [38]. Du et al. [39] claimed that in long-standing CRVO eyes, there was no significant change in SFCT after anti-VEGF treatment. This may be related to the different durations of RVO onset and follow-up times in the included cases. We treated all patients with a 3+PRN regimen. For both CRVO and BRVO, BCVA and CRT in the affected eyes were significantly improved after three injections. Choroidal thickness, LA, SA, TCA, and CVI all decreased after the first and second injections and remained relatively stable thereafter. We speculate that the main reason for the rapid thinning of choroidal thickness after intravitreal injection of ranibizumab is that ranibizumab can penetrate the retina and reach the choroid. While improving retinal barrier function and reducing retinal capillary permeability, it also reduces choroidal vascular permeability, thereby reducing choroidal thickness. At the same time, we compared various indicators of different types of RVO macular edema. We found that regardless of the type of RVO-ME patient, BCVA and CRT in the affected eyes were significantly improved after multiple injections. Choroidal thickness, LA, SA, and TCA all decreased after the first injection and remained relatively stable after the second injection. This indicates that anti-VEGF treatment can reduce choroidal blood flow perfusion. The CVI of CME and mixed types also decreased after the first injection and remained relatively stable after the second injection. However, the changes in CVI of DRT type after multiple injections were not statistically significant. The specific reasons are still unclear and may be related to the relatively small number of DRT types in this study, which may lead to biased results. At the same time, we also compared the indicators of the three types of macular edema after anti-VEGF treatment. We found that there were no statistically significant differences in choroidal indicators among the three types of macular edema. This suggests that changes in choroidal vasculature may be unrelated to the type of edema.

After repeated anti-VEGF treatment in the acute phase of RVO, macular edema disappears. Over time, collateral circulation gradually opens, and the condition enters a stable phase. We retrospectively reviewed the data of 70 RVO patients in the stable phase who had macular edema resolution and had stopped injection for at least six months. We analyzed their choroidal indicators and observed the dynamic changes in choroidal indicators after multiple ranibizumab treatments during the acute phase of RVO and at least six months after stopping injection in the stable phase. We found that after entering the stable phase and stopping injection, CRT in RVO patients remained relatively stable compared to the last injection, but BCVA further improved compared to the last injection. This suggests that photoreceptor affected by macular edema may retain functional recovery potential following edema resolution.

Regardless of the type of RVO or RVO-ME, choroidal thickness gradually decreased after anti-VEGF treatment and remained relatively stable thereafter. LA, SA, TCA, and CVI gradually decreased after anti-VEGF injection, with the downward trend continuing for at least six months after the last injection. This shows that after treatment in the acute phase of RVO, the intraocular VEGF levels decreased, choroidal vascular leakage was reduced, and choroidal vasculature was further remodeled.

In addition, we found that males are more likely to experience changes in CVI than females, and BRVO is more likely to cause changes in CVI than CRVO. Other indicators such as age, type of macular edema, baseline visual acuity, and baseline CRT showed no significant correlation with changes in CVI. The specific mechanisms of these factors affecting CVI need to be further explored.

In this study, we chose CVI as our research indicator, which has certain advantages. It is less affected by physiological factors and has less variability compared to traditional choroidal thickness. By measuring the areas of LA and SA, it can assess the choroidal structure and perfusion, helping to better understand the pathological mechanisms of retinal diseases and evaluate the response to treatment. Although CVI is recognized as a relatively reliable and promising non-invasive tool for studying choroidal vasculature, it still has some limitations in clinical use. First, high-quality, clear EDI-OCT scan images are required; otherwise, the reliability of CVI is poor. Second, the measurement area boundaries of CVI need to be manually drawn, which is inevitably subject to subjective factors. Third, the EDI-OCT images obtained need to be processed using Image J software, which requires some knowledge of the software and is relatively complex and time-consuming. To avoid these limitations, we excluded all OCT images with unclear choroidal imaging. In addition to excluding images that were unclear due to severe cataracts or vitreous hemorrhage, we also excluded those with unclear choroidal images due to extensive retinal hemorrhage or high macular edema in the acute phase of RVO. All images were manually drawn by the same experienced physician to ensure the accuracy of the data in this study and minimize bias caused by human factors. At the same time, CVI cannot reflect the blood flow perfusion of specific layers of the choroidal capillaries, medium vessels, and large vessels; it can only reflect the overall choroidal blood flow perfusion. Therefore, some scholars have begun to explore the use of the H/C ratio (Haller layer/choroidal thickness) to distinguish the ability of RVO eyes from healthy eyes. Aribas YK et al. [16] believe that the intravascular pressure in the large vessels of the choroidal Haller layer is the highest. Therefore, choroidal congestion will not cause it to shrink, and the increase in Haller layer thickness will further compress the smaller vessels, namely the Sattler layer/choroidal complex. They found that changes in CVI and H/C ratio were accompanied by a decrease in choroidal capillary blood flow density. Nowadays, the measurement tools of some OCT systems can easily calculate the H/C ratio. Therefore, the H/C ratio seems to be easier to implement in clinical practice than CVI and needs further exploration and validation. At the same time, we also look forward to the progress of computer image processing methods and the driving of artificial intelligence technology to make the application of CVI more convenient and mature in the future [40].

Innovations and Limitations of This Study: Currently, in clinical practice, the choroidal morphology in RVO patients is mostly represented by the subfoveal choroidal thickness. In addition to the subfoveal choroidal thickness, our study measured the choroidal thickness 1500μm nasal and temporal to the macular fovea to calculate the mean choroidal thickness in the macular center area. Multiple measurements can improve the accuracy of choroidal thickness. Moreover, we used the CVI index to assess the structure and morphology of the choroid, which can significantly reduce bias caused by variations in choroidal thickness.A few studies have reported changes in choroidal thickness in RVO patients after anti-VEGF treatment, mostly focusing on the results of a single injection, with inconsistent findings. We compared the choroidal thickness and CVI of different types of RVO and also examined the changes in choroidal thickness and CVI after multiple anti-VEGF treatments, aiming to provide more clinical references for understanding the choroidal data of different types of RVO patients. We compared the changes in choroidal indicators during the acute and stable phases of RVO, bringing new insights into assessing the changes in choroidal vasculature caused by RVO at different stages of the disease. Limitations of This Study: First, although we had the same doctor measure the choroidal thickness, there is always potential bias in manual measurements. We look forward to the development of automated software for more objective and precise assessments in the future. Second, choroidal thickness exhibits certain rhythmicity, with differences when measured at different times of the day. As this was a retrospective study, we did not perform OCT measurements on all patients during the same time period, which may introduce some bias. Choroidal thickness is influenced by factors such as age, time of day, and axial length, making it difficult to ensure completely consistent measurement conditions. To minimize bias in choroidal measurements, we excluded patients with refractive error equivalent spherical ≥±6.0D, axial length >26mm or <22mm during enrollment. We also introduced the choroidal vascularity index (CVI) as a new indicator to assess choroidal perfusion. CVI is less affected by the above factors and can serve as a new observational indicator to compensate for the limitations. In the future, we plan to conduct multicenter prospective studies to provide more reliable reference data for clinical practice.

## Conclusions

Retinal vein occlusion (RVO) not only affects the retinal structure but also has an impact on the morphology and blood flow perfusion of the choroid. The choroidal vessels in RVO eyes are dilated, with higher choroidal thickness and CVI compared to non-occluded eyes. CRVO has greater choroidal thickness than BRVO, but CVI values do not differ based on the type of RVO or the type of RVO macular edema. After treatment in the acute phase of RVO, choroidal thickness, LA, SA, TCA, and CVI all decrease and remain relatively stable thereafter, indicating ongoing remodeling of the choroidal vasculature. CVI shows no significant correlation with age, visual acuity, type of macular edema, or central retinal thickness, making it a reliable indicator for assessing choroidal blood flow perfusion. Baseline choroidal thickness and CVI can serve as predictive factors for improvement in BCVA and CRT after anti-VEGF treatment.

## Data Availability

All relevant data are within the manuscript and its Supporting Information files.

## List of abbreviations

RVO: Retinal Vein Occlusion
CRVO: Central Retinal Vein Occlusion
BRVO: Branch Retinal Vein Occlusion
CSC: Central Serous Chorioretinopathy
PCV: Polypoidal Choroidal Vasculopathy
CNV: Choroidal Neovascularization
AMD: Age-related Macular Degeneration
DR: Diabetic Retinopathy
DME: Diabetic Macular Edema
PDT: Photodynamic Therapy
ME: Macular Edema
CRT: Central Retina Thickness
SFCT: Subfoveal Choroidal Thickness
CT N1.5mm: Nasal Choroidal Thickness at 1.5mm of macula
CT T1.5mm: Temporal Choroidal Thickness at 1.5mm of macula
CT mean: Mean Macular Thickness
LA: Luminal Area
SA: Stromal Area
TCA: Total Choroidal Area
CVI: choroidal vascularity index
EDI: Enhanced Depth Imaging
anti-VEGF: anti-Vascular Endothelial GrowthFactor
BCVA: Best-Corrected Visual Acuity
IOP: Intraocular Pressure
OCT: Optical Coherence Tomography
DRT: Diffuse Retinal Thickening
CME: Cystoid Macular Edema
SRD: Serous Retinal Detachment
CRT: Central Retinal Thickness

## Author’s Contributions

Yuanyuan Qi conducted data curation, formal analysis, and contributed to drafting, reviewing, and editing the manuscript; Jiayang Xu contributed to data curation and investigation; Daniel Hillarion Scotland conducted reviewing and editing; Chao Zhang contributed to methodology. All authors read and approved the final manuscript.

## Availability of data and materials

All relevant data are within the manuscript and its Supporting Information files. Data and materials can be asked from the correspondence author by Email.

## Ethics approval and consent to participate

This study was approved by the institutional review board of Dalian No.3 hospital’s ethics review committee (2023-145-001). It was conducted in accordance with the principles of the Declaration of Helsinki.

## Consent for publication

Not applicable.

## Acknowledgments

Not applicable.

